# Early microbiome and metabolome signatures in pediatric patients undergoing allogeneic hematopoietic cell transplantation

**DOI:** 10.1101/2021.06.08.21258499

**Authors:** Caitlin W. Elgarten, Ceylan Tanes, Jung-jin Lee, Lara A. Danziger-Isakov, Michael S. Grimley, Michael Green, Marian G. Michaels, Jessie L. Barnum, Monica I. Ardura, Jeffery J. Auletta, Jesse Blumenstock, Alix E. Seif, Kyle L. Bittinger, Brian T. Fisher

## Abstract

**Background:** The contribution of the gastrointestinal tract microbiome to outcomes after allogeneic hematopoietic cell transplantation (HCT) is increasingly recognized. Investigations of larger pediatric cohorts aimed at defining the microbiome state and associated metabolic patterns pre-transplant are needed.

**Methods:** We sought to describe the pre-transplant stool microbiome in pediatric allogenic HCT patients at four centers. We performed shotgun metagenomic sequencing and untargeted metabolic profiling on pre-transplant stool samples. Samples were compared with normal age-matched controls and by clinical characteristics. We then explored associations between stool microbiome measurements and metabolite concentrations.

**Results:** We profiled stool samples from 88 pediatric allogeneic HCT patients, a median of 4 days before transplant. Pre-transplant stool samples differed from healthy controls based on indices of alpha diversity and in the proportional abundance of specific taxa and bacterial genes. Relative to stool from healthy patients, samples from HCT patients had decreased proportion of Bacteroides, Ruminococcaeae and genes involved in butyrate production, but were enriched for gammaproteobacterial species. No systematic differences in stool microbiome or metabolomic profiles by age, transplant indication or hospital were noted. Stool metabolites demonstrated strong correlations with microbiome composition.

**Discussion:** Stool samples from pediatric allogeneic HCT patients demonstrate substantial dysbiosis early in the transplant course. As microbiome disruptions associate with adverse transplant outcomes, pediatric-specific analyses examining longitudinal microbiome and metabolome change are imperative to identify causal associations and to inform rational design of interventions.

## INTRODUCTION

The contribution of the human microbiota to human physiology, health, and disease is increasingly recognized. Recent studies have associated alterations of the intestinal microbiota with an increased or decreased risk of important outcomes after allogeneic HCT, including bloodstream infection,^1,2^ GVHD,^3-6^ pulmonary complications,^7^ immune reconstitution,^8^ relapse^9^ and all-cause mortality.^10-12^ While evidence regarding these associations is accumulating among adult allogeneic HCT recipients, far less is known in the pediatric setting. Healthy children demonstrate microbiome community structure and diversity that evolve with the immune system in an age-dependent manner.^13-15^ As this evolution of the microbiome differs from adults, data on adult microbiome phenotypes and their association with specific post-HCT outcomes are likely not generalizable to children.^13^

To date, studies of the microbiome specific to pediatric HCT recipients have been limited by relatively small patient numbers.^6,16,17^ Further, the majority of studies have assessed the microbiome at the time of neutrophil engraftment post-HCT.^2,10,18-20^ Fewer studies have considered the pre-transplant microbiome with conflicting results regarding its composition.^4,5,12,21^ Better understanding of the microbiome at a pre-transplant timepoint may provide insights into opportunities to develop clinically relevant microbiome assays to predict, diagnose or prevent downstream complications. Similarly, measurement of the metabolome has emerged as a mechanism to connect environmental exposures, the gastrointestinal microbiota and post-transplant outcomes. Metabolic profiles in the blood and intestinal tissue associate with the later development of adverse post-transplant outcomes,^22-25^ but stool metabolic patterns, particularly in pediatrics, have not been well described.

Given the limited knowledge in microbiome and metabolome studies in pediatric allogeneic HCT, investigations with larger pediatric cohorts aimed at determining the microbiome state and associated metabolic patterns prior to HCT are needed. The goal of this study was to characterize the pre-HCT microbiome and metabolome of children across four centers and to identify connections between microorganisms and biochemicals present in the gut. We also explored the impact of clinical variables such as indication for transplant, hospital location and age at transplant on microbiome and metabolome composition. Foundational data from this descriptive multicenter observational study will serve to generate hypotheses and inform study design of future interventional studies.

## METHODS

### Patients, specimens, and clinical data collection

This was an ancillary study of patients prospectively enrolled to the Adenovirus in Pediatric Transplant Study (AdOPT) at four pediatric transplant centers from October 2017 to September 2018 (DMID Protocol Number 160097). AdOPT aims to assess the impact of adenovirus infection in the peri-HCT period. All pediatric patients undergoing an allogeneic HCT for any indication in the time period were eligible for inclusion in the AdOPT cohort. Subjects were enrolled prior to transplant and had a baseline stool specimen obtained between day minus seven and the day of transplant. The stool specimen was obtained as part of the AdOPT study to establish presence or absence of adenovirus shedding in the gastrointestinal tract. After processing for adenovirus testing, remnant stool specimens were stored at −80 C. These remnant specimens were retrieved for the microbiome and metabolome analyses in this study.

Demographic, clinical and laboratory data were manually abstracted from the electronic medical record into a REDCap™ database. Indication for transplant was classified as malignancy, bone marrow failure syndromes, immune deficiency/immune dysregulation syndromes (including primary immunodeficiencies and hemophagocytic lymphohistiocytosis) and other (including hemoglobinopathies and metabolic diseases). Subjects with stool sample remaining after adenovirus analyses who had provided written informed consent for secondary use of research materials were included in these analyses. This study protocol was approved by the CHOP Institutional Review Board, the IRB of record for all centers.

Stool samples from healthy controls that were previously collected and sequenced^26,27^ were employed as comparators. These were healthy pediatric volunteers without significant medical illness, diarrhea, history of antibiotic use in the past six months, use of immunosuppressive medications or recent travel.

### Microbial DNA extraction, sequencing, and bioinformatics

DNA was extracted from whole stool and sequenced using the Illumina HiSeq paired-end method. Taxonomic assignment was performed using Sunbeam, a user-extendable bioinformatics pipeline developed at the CHOP Microbiome Center.^28^ Quality control steps were performed to remove host-derived sequences and reads of low sequence complexity. Bacterial abundance was estimated using Kraken;^29^ abundance of bacterial gene orthologs was estimated using the KEGG database,^30^ as well as to curated databases of genes involved in butyrate production, polysaccharide utilization, and secondary bile acid production.^31,32^ Samples were summarized using Shannon diversity index and richness as measures of alpha diversity and dysbiosis defined as the distance the centroid of age-matched healthy samples.

### Metabolome analyses

Untargeted metabolomic profile analyses were performed by Metabolon Inc (Durham, NC, USA) as described previously.^33^ Samples were prepared using the automated MicroLabSTAR system (Hamilton Company, Bonaduz, Switzerland) and extracted using Metabolon ‘s standard solvent extraction method. Small molecules were dissociated from proteins and proteins were removed. Samples then underwent two separate reverse phase (RP)/ultra-performance liquid chromatography tandem mass spectrometry (UPLC-MS/MS, Waters ACQUITY, Milford, MA, USA) methods with positive ion mode electrospray ionization (ESI, Thermo Scientific, Waltham, MA, USA) RP/UPLC-MS/MS with negative ion mode ESI, and hydrophilic interaction liquid chromatography (HILIC)/UPLC-MS with negative ion mode ESI. Metabolites were identified by comparison to a library maintained by Metabolon Inc. based on retention time/index, mass to charge ratio, and chromatographic data, and quantified using area-under-the-curve.

### Statistical analyses

Comparisons were made between transplant patients and healthy controls, and then among transplant patients by indication for transplant, hospital location, and age group. As exposures that could impact the microbiome composition accumulate over the days that the preparative regimen is administered, we also examined microbiome indices by day of stool collection. Pairwise Wilcoxon rank sum tests were used to compare alpha diversity between groups using normalized numbers of microbial reads. Bray-Curtis and Jaccard distance were used to construct Principal Coordinates Analysis graphs. Random Forest analysis (RFA) was used to discern how well individuals could be classified into each group based on genus-level relative abundances.

To determine the relationships between microbiome measurements and metabolomic profiles, correlations between the log transformation of metabolite abundance and dysbiosis were calculated using linear regression. In addition, alpha diversity and butyrate levels were evaluated using Pearson correlation coefficient and linear regression, based on data from adult cohorts identifying an association between these values specifically.^34,35^ Spearman correlation was calculated for the metabolites that are present in at least 80% of the samples and the bacteria that have at least 1% mean relative abundance across all samples.

All p-values were corrected for multiple comparisons using the Benjamini-Hochberg methods. A false discovery rate (FDR) of 5% was used. Calculation of microbiome measures and statistical analyses were performed with R, version 3.4.2 (R Development Core Team, Vienna, Austria).

## RESULTS

Pre-transplant stool samples were obtained at a median of 4 days before transplant from 88 allogeneic HCT recipients at four pediatric centers. Cohort characteristics are shown in Table 1. The median age of patients was 6.2 years (range, 0.33–17.9 years) and the most common indications for HCT were malignancy (n=35, 39.8%) and immunodeficiency/dysregulation (n=33, 37.5%). Microbiome results of stool specimens previously processed from 52 healthy controls were included in these analyses. The median age of the healthy controls was 5.6 years (range, 0.33–17.6 years).

**TABLE 1.**
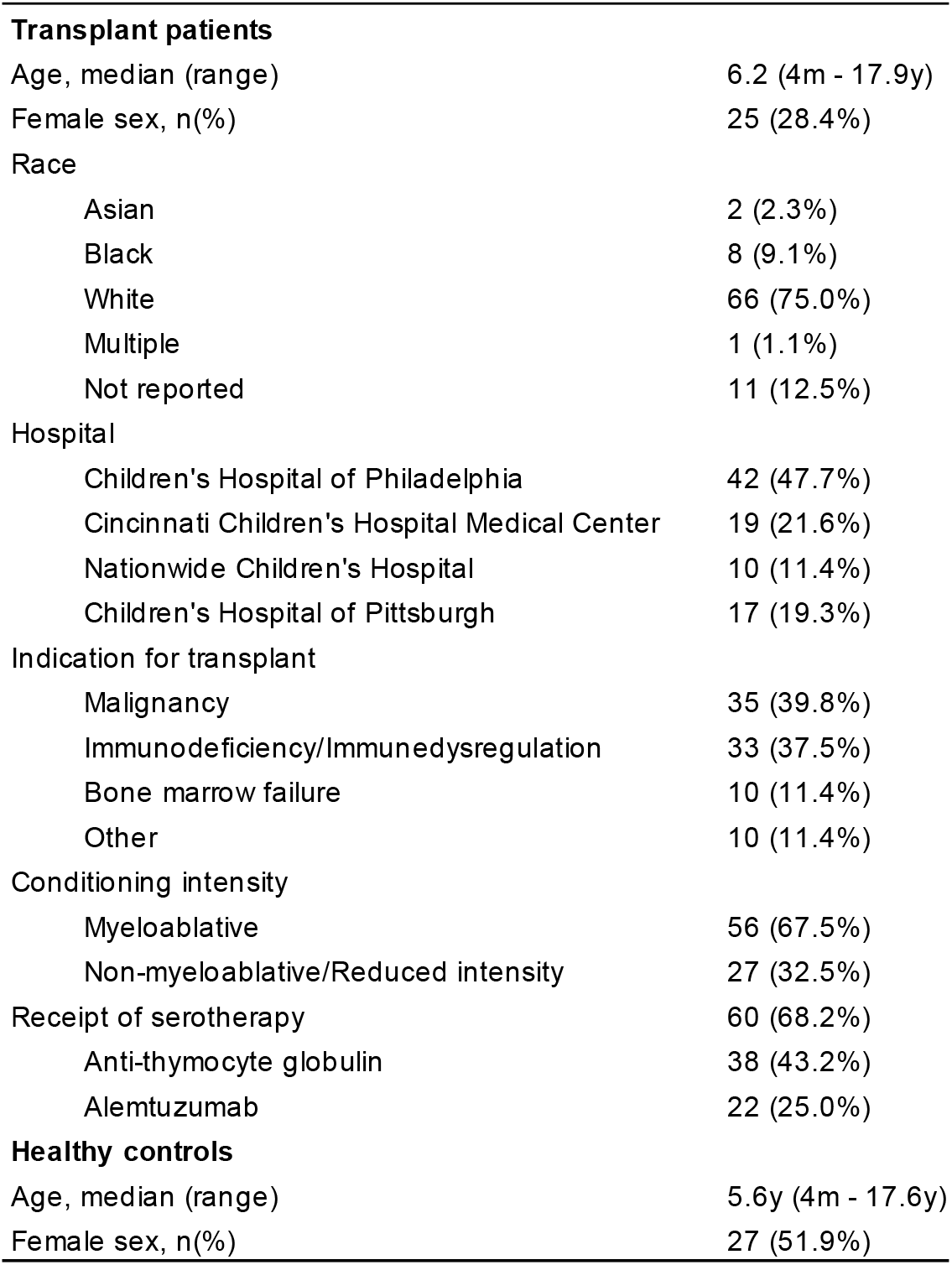
Cohort Characteristics:

### Comparison of taxa between HCT patients and age-similar healthy children

We compared the gut microbiome composition of patients in the allogeneic HCT cohort with the composition of healthy, age-similar children. Compared with healthy individuals, pre-HCT patients had significantly lower microbial diversity according to both richness and Shannon diversity index (Figure 1A). Beta diversity plots revealed differences between HCT patients and healthy controls (Figure 1B). The variation was driven primarily by a decrease in median abundance of certain genera from the Bacteroidetes and Clostridiales order in the HCT patients, including a decreased proportion of Bacteroides, Ruminococcaeae and *Akkermansia muciniphilia* (FDR controlled p(q)<0.05). Relative to healthy children, samples from HCT patients were enriched for gammaproteobacterial species (e.g. *Klebsiella* species, *Escherichia coli)* and *Enterococcus faecalis* (Figure 1C). A heatmap of bacterial taxa at the genus level is shown in supplementary Figure 1.

**FIGURE 1:**
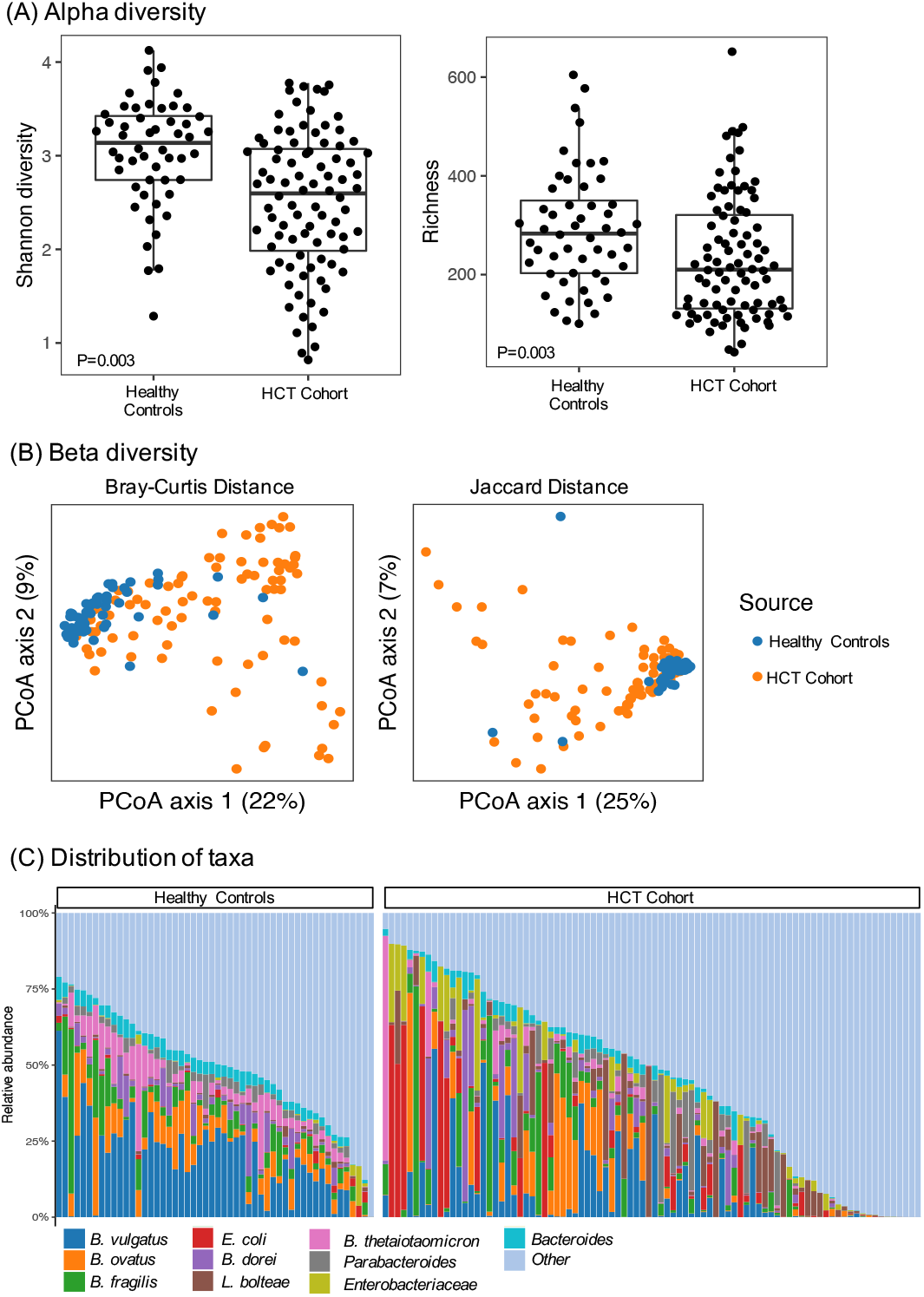
Taxonomic composition of samples from pre-HCT patients compared to age-similar normal healthy. (A) Alpha diversity comparisons by Shannon diversity (left panel) and richness (right panel). Wilcoxon signed rank test p values shown. (B) Visualization of compositional profiles among HCT patients and healthy controls using principal coordinate analysis. The PCoA was conducted based on Bray-Curtis distance (left panel) and Jaccard distance (right panel). Each point represents a single stool sample. The more similar the samples are, the closer together they appear. (C) Distribution of relative abundance of ten most abundant taxa in HCT cohort and healthy controls.

### Comparison of gene abundance between HCT patient and controls

Based on gene abundance, we again saw clear separation between HCT patients and healthy controls based on Bray-Curtis distances. When we examined genes involved in butyrate metabolism specifically, both butyryl-CoA dehydrogenase and butyrate kinase were decreased in abundance compared with healthy controls (Figure 2).

**FIGURE 2:**
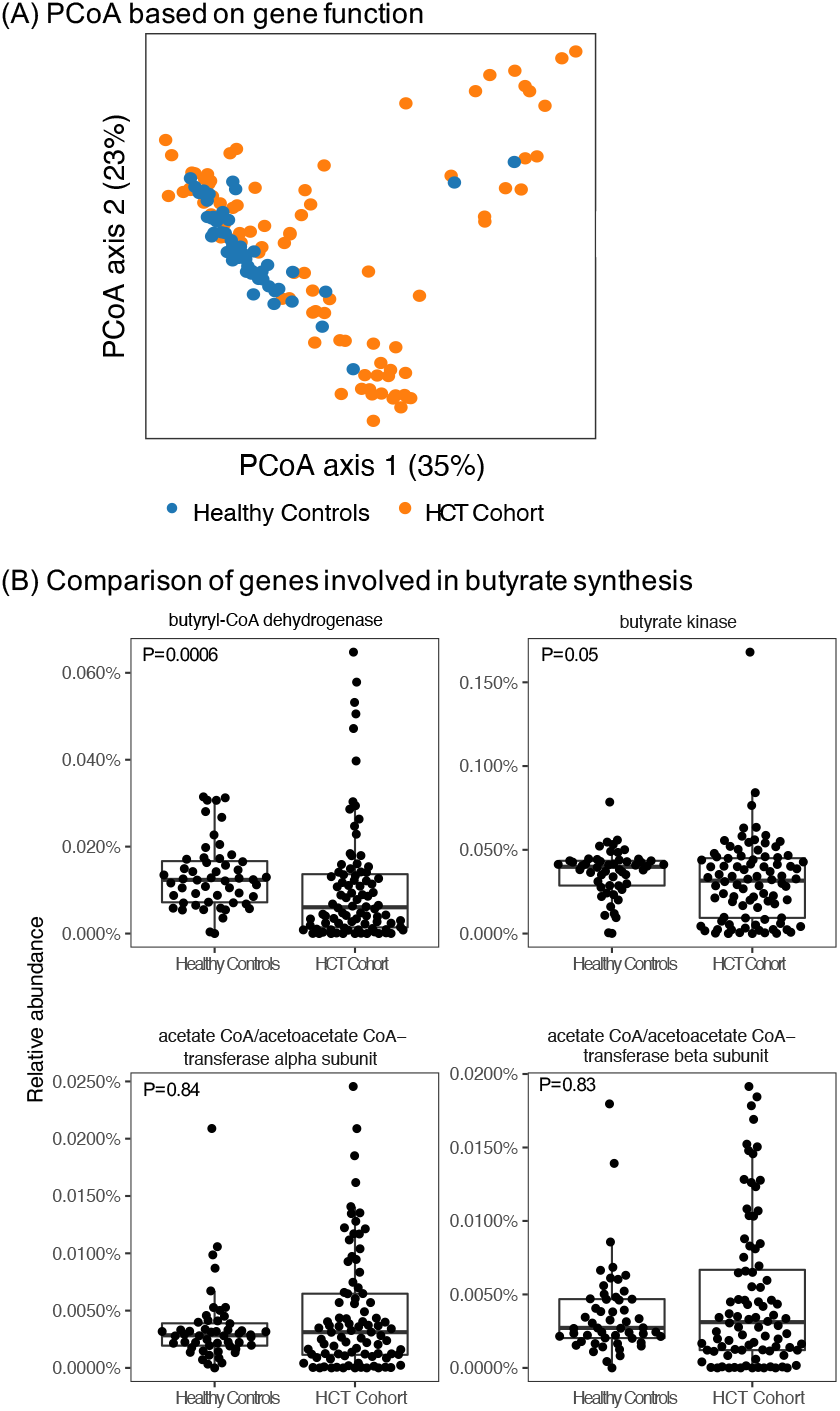
Gene abundance of samples from pre-HCT compared to age-similar healthy controls. (A) PCoA conducted based on Bray-Curtis distance calculated based on gene function and demonstrating varying compositional profiles between the pre-HCT cohort and age-similar healthy controls. (B) Comparison of abundance of genes involved in butyrate pathways. Wilcoxon signed rank test p values shown.

### Microbial and gene function composition by sub-groups

Within the cohort of allogeneic HCT recipients the pre-transplant microbial composition was compared by transplant indication, age, and hospital location (Figure 3). Clustering of all samples using the Bray-Curtis dissimilarity metric demonstrated that within clinical characteristics samples were heterogenous and departed from the cluster of healthy controls, but none of these groups separated the transplant patients from one another. We identified no other systematic differences in alpha diversity, taxon or gene abundance by any of these group variables. By random forest analysis, the genus level microbiome composition did not discriminate between transplant indication (error rate 54-100% for each indication) or hospital location transplant (error rate 11-100% for each hospital).

**FIGURE 3:**
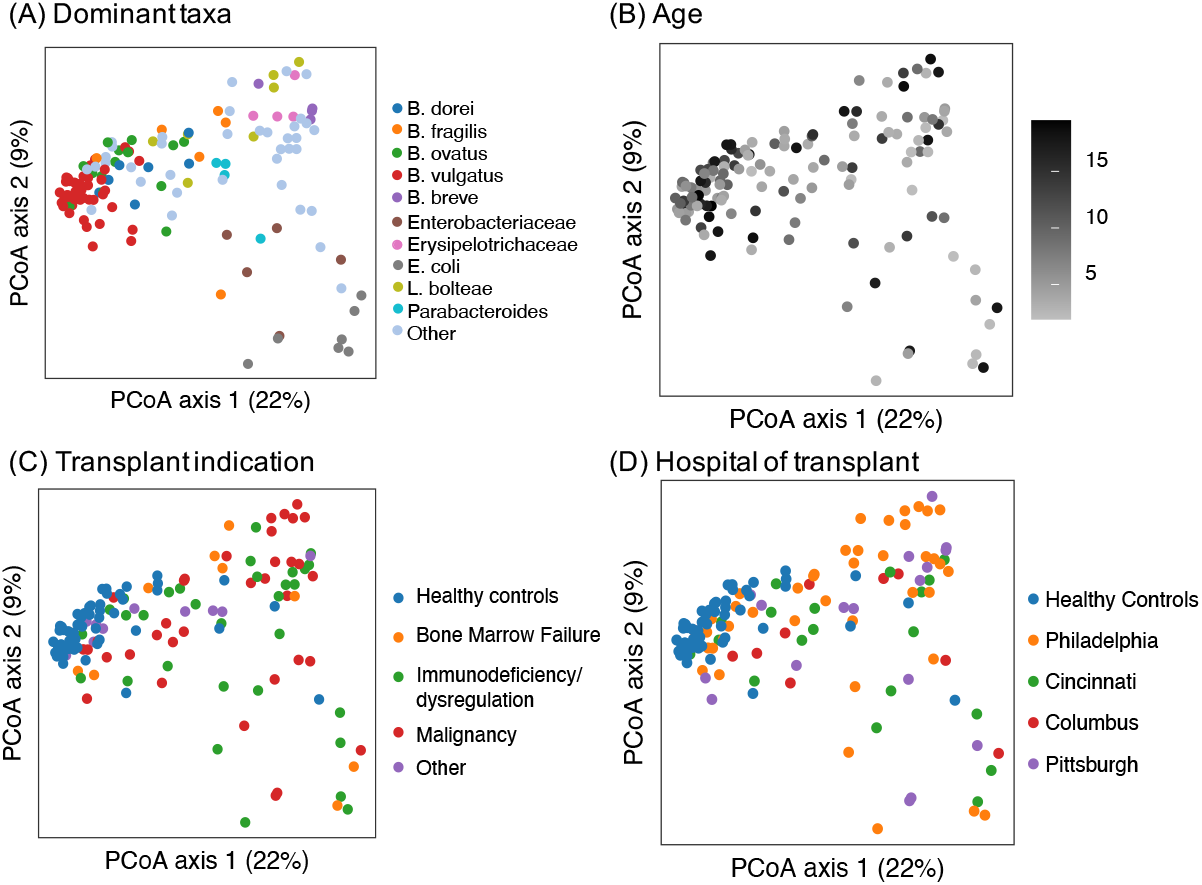
Assessment of clinical characteristics impacting microbiome configuration. Samples from the pre-HCT cohort and healthy controls. In panel (A) the samples are coded by most common taxa. The samples do not show any separation by age (B), transplant indication (C) or hospital of transplant (D).

### Microbiome-derived metabolite composition

Untargeted metabolomic profiling identified 1,226 informative biochemicals. These metabolites could be divided into the eight main categories: amino acids (254), peptides (43), carbohydrates (42), energy metabolism (14), lipids (459), nucleotides (70), cofactors/vitamins (63), and xenobiotics (276). Clinically relevant metabolites demonstrated substantial heterogeneity between samples. No individual biochemicals reached our FDR adjusted threshold for significance when compared by clinical sub-groups.

### Cross-sectional associations between metagenomic data and metabolite production

In our cohort, stool butyrate levels did not correlate with Shannon diversity (r=0.07, p=0.42), and only a slight correlation was noted between stool indole levels and Shannon diversity (r=0.22, p=0.04). However, when we examined the association between biochemicals and dysbiosis, 448 were significantly associated by linear regression with an FDR p<0.05. The highly correlated metabolites are shown in Figure 4; secondary bile acids, as well as some amino acid derived metabolites were strongly associated with dysbiosis. Examination of the overall network structure demonstrating correlations between bacterial genera and metabolites is presented in Figure 5.

**FIGURE 4:**
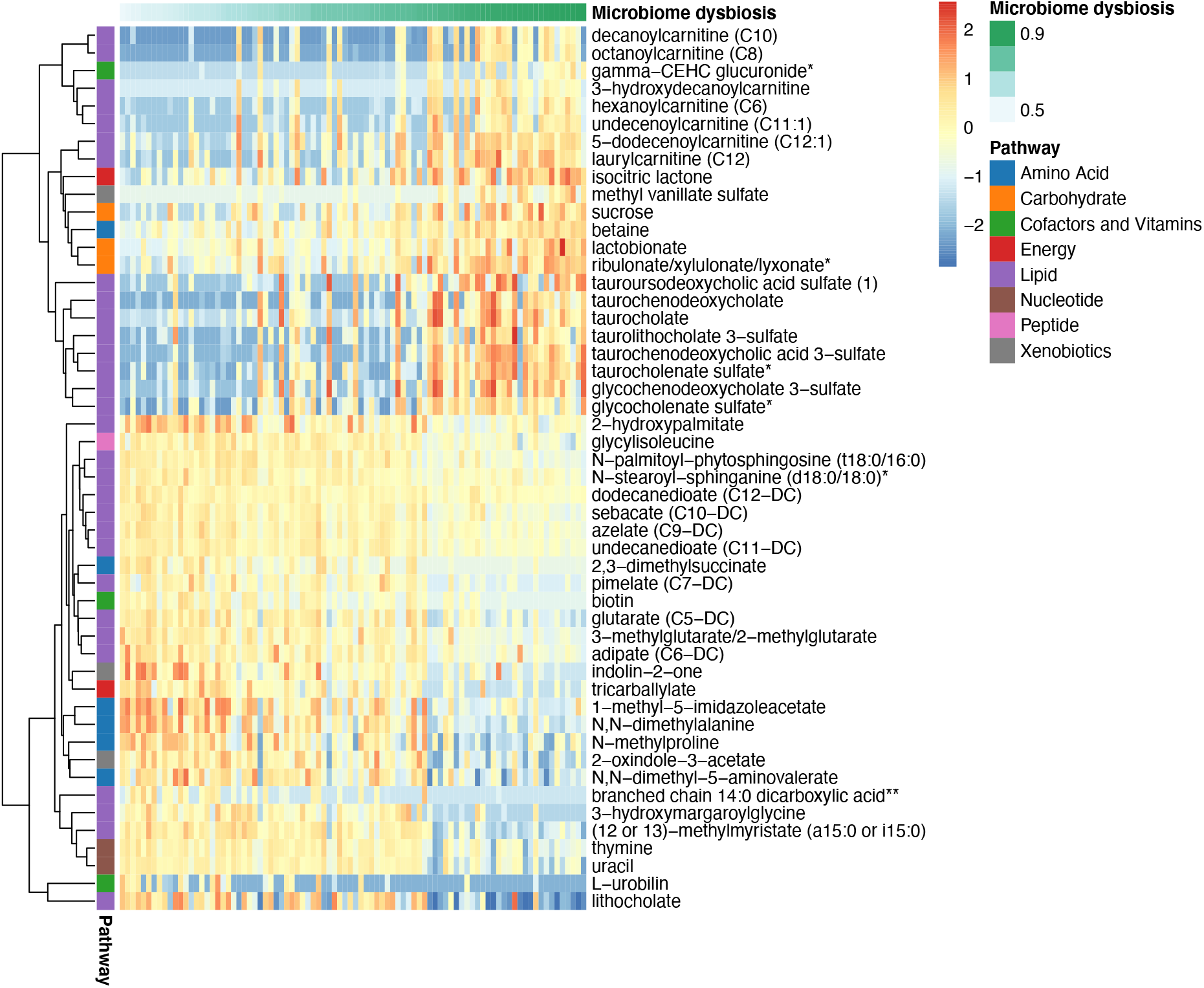
Heatmap of stool metabolites sorted by dysbiosis. For samples from the pre-HCT cohort, linear regression was used to associate metabolite abundance with dysbiosis and the most correlated metabolites are shown here, clustered by Manhattan distance.

**FIGURE 5:**
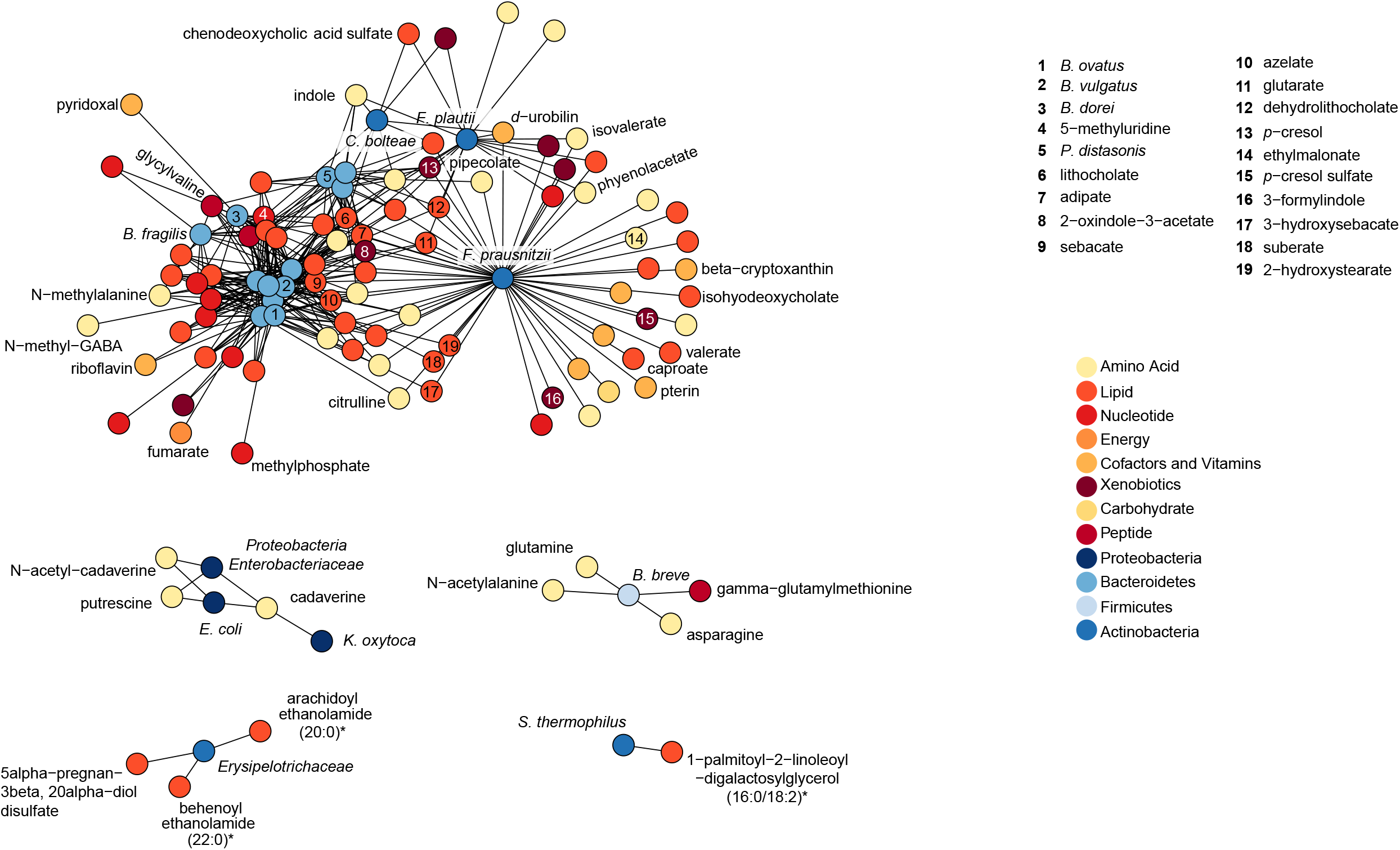
Network analysis demonstrating relationships between bacterial taxa and metabolites. Spearman correlation was calculated for the metabolites that are present in at least 80% of the samples and the bacteria that have at least 1% mean relative abundance across all samples. P values were adjusted for false discovery rate using Benjamini-Hochberg method. The resulting positive correlations with FDR adjusted p value <0.01 are shown.

## DISCUSSION

In this descriptive study, we employed metagenomic sequencing and untargeted metabolomic profiling to characterize stool samples of children undergoing allogeneic HCT at four US centers in order to enhance our understanding of the gut microbiome of children in the immediate pre-transplant period. Collectively, the results suggest that disturbance of the microbiome in children begins before the transplant and well before engraftment. Pre-transplant stool specimens demonstrated a decrease in microbial diversity and differences in microbial composition compared with healthy age-matched controls. These results are consistent with data from adult studies that found lower pre-transplant bacterial diversity and different taxa abundance compared to both healthy controls at the same center and participants of the human microbiome project.^12,21,35^

We sought to identify clinical characteristics that would be associated with microbiome or metabolome composition in this patient population; however, analyses of age, hospital of transplant, and indication for transplant did not identify microbiota composition or metabolic phenotypes associated with these variables. Previous work by Biagi et al. has demonstrated that dysbiosis in the peri-transplant period occurs in an individual-specific manner, which is consistent with the “Anna Karenina principle”: that each dysbiotic microbiome configuration is uniquely dysbiotic.^13,16,37^ Our cross-sectional data suggest that the microbiome of children are perturbed extensively enough early in the pre-transplant period to override any age- or hospital-specific microbiota signatures that may exist in healthy patients by the time the HCT takes place. This likely reflects exposures during their preparative regimens (e.g. prophylactic antimicrobials, conditioning agents), as well as the extensive medical contact children undergoing allogeneic HCT have prior to their transplant admission which can lead to persistent changes in the commensal bacterial composition.^38-44^ Additional research is needed to determine the major

Comparison of taxa and gene abundance between our allogeneic HCT recipients and healthy controls provides specific details regarding how the microbiome is perturbed for children in the pre-HCT period and suggests that these perturbations may be clinically meaningful. To date most associations of dysbiosis with post-transplant outcomes have been based on stool specimens collected later in the transplant process, particularly at the time of neutrophil engraftment. While there are some data in adults associating dysbiosis earlier in the transplant period with outcomes after HCT ^12,34^, there have been limited data for children. Prior small pediatric studies have yielded conflicting results. A cohort of six children undergoing HCT for ALL had decreased alpha diversity compared to healthy children^36^ whereas a report of ten children undergoing HCT for malignant and non-malignant indications reported that the microbial taxa distribution approximated profiles found in healthy controls.^5^ Data from our larger cohort supports that children undergoing conditioning for allogeneic HCT have an already perturbed microbiome in the peri-transplant period.

This foundational information raises the possibility for an opportunity to manipulate the pediatric microbiome in the pre-transplant period to alter downstream risk of outcomes such as bacteremia,^2^ pulmonary complications,^7^ GVHD^4,6,11^, and mortality.^10^ Further investigation is necessary to understand how these pre-transplant perturbations may be mechanistically associated with subsequent negative outcomes after transplant. However, these data suggest that any attempt to associate changes in the microbiome with post-HCT outcomes needs to consider the trajectory of microbiome changes starting with sampling well antecedent to the transplant.

Stool metabolites may indicate potential pathways by which the microbiome modulates associations between the microbiome and post-transplant outcomes.^45-47^ Metabolites that have been identified as potentially clinically relevant in other cohorts, including short chain fatty acids, amino acid-derived metabolites (tryptophan, tyrosine, choline), riboflavin-derived metabolites and bile acids demonstrated substantial variability between pre-transplant samples.^48^ We examined previously described adult associations between microbiome diversity and the metabolome. In adults, stool butyrate and indol levels have been associated with microbial disruption,^34,35^ however, we only identified a minimal association between stool indol levels and increased diversity and no association between butyrate levels and diversity.

Instead, metabolite concentrations, especially secondary bile acids (e.g. tauroursodeoxycholic acid), were highly correlated with dysbiosis. Bile acid malabsorption by the terminal ilium has been identified as a biomarker for GVHD or even a contributor to gastrointestinal GVHD pathogenesis.^22,49^ Moreover, our network analysis demonstrated related clusters of biochemicals around Bacteroides, Parabacteroids, and *Faecalibacterium prausnitzii;* whereas, there were distinct clusters of biochemicals anchored by Proteobacteria, *Erysipelotrichaceae*, as well as *Bifidbacterium breve* and *Streptococcus thermophilus*. This network structure demonstrates important associations between both organisms and metabolites that have been previously identified as inflammatory or immunoregulatory.^48^ The heterogeneity of the microbiome and metabolomic layouts early in the transplant course suggests that this may be a potential arena for further exploration within pediatric transplantation.

This study was limited in its cross-sectional nature. Comparisons identified in this sample are hypothesis-generating but longitudinal characterization of the microbiome and microbiome-derived metabolites are needed to derive causal inferences about drivers and implications of microbiome change. In addition, these samples were collected during the admission for HCT, and therefore the identified dysbiosis reflects exposures that accumulate during the preparative regimen, as well as the baseline state prior to admission. Importantly, our analyses did not identify any systematic difference in microbiome composition by day of collection during this time period. It will be imperative that future efforts capture stool samples even further antecedent to the transplant. Finally, although this is a large pediatric cohort, there was substantial heterogeneity with regards to patient and transplant characteristics, and numbers limited the ability to perform subgroup analyses. However, the finding that the microbiome did not vary systematically by hospital location suggests that combining patients across institutions is reasonable to establish larger cohorts that would allow for nuanced assessment of the microbiome in this patient population.

In conclusion, we found that the intestinal microbiomes of children admitted for HCT were significantly deranged compared to normal controls, and highly correlated with metabolomic signatures, including specific changes that have been previously associated with poor clinical outcomes. Although the implications of these pre-transplant perturbations have not yet been definitively established in pediatrics, their presence early in the transplant period makes further investigation of causal association with outcomes imperative. Importantly, if longitudinal studies confirm a mechanistic link to post-transplant outcomes, this time point before significant mucositis or neutropenia offers an opportunity for restoration of normal microbiota. Given the sample size limitations inherent in studies involving pediatric transplant recipients, and the growing evidence that the microbiome does not function identically in children and adults,^50^ additional multicenter collaborative efforts are necessary to further clarify the role of the microbiome starting from prior to transplant in determining post-transplant outcomes.

## Data Availability

The data that support the findings of this study are available from the corresponding author upon reasonable request.

## ACKNOWLEDGEMENTS

This work was supported by grants from the Precious Jules Foundation and Hematologic Malignancy section of the Center for Childhood Cancer Research at CHOP (CE) and a contract (N01AI2014028) from NIH/NIAID (BF). Craig Boge, MPH and Sydney Schuster, MPH were instrumental in accomplishing this work.

## REFERENCES

1. Taur Y, Pamer EG. Microbiome mediation of infections in the cancer setting. Genome Med. 2016;8(1):40.

2. Taur Y, Xavier JB, Lipuma L, et al. Intestinal domination and the risk of bacteremia in patients undergoing allogeneic hematopoietic stem cell transplantation. Clin Infect Dis. 2012;55(7):905–914.

3. Jenq RR, Taur Y, Devlin SM, et al. Intestinal Blautia Is Associated with Reduced Death from Graft-versus-Host Disease. Biol Blood Marrow Transplant. 2015;21(8):1373–1383.

4. Doki N, Suyama M, Sasajima S, et al. Clinical impact of pre-transplant gut microbial diversity on outcomes of allogeneic hematopoietic stem cell transplantation. Ann Hematol. 2017;96(9):1517–1523.

5. Biagi E, Zama D, Nastasi C, et al. Gut microbiota trajectory in pediatric patients undergoing hematopoietic SCT. Bone Marrow Transplant. 2015;50(7):992–998.

6. Tanaka JS, Young RR, Heston SM, et al. Anaerobic Antibiotics and the Risk of Graft-Versus-Host Disease after Allogeneic Hematopoietic Stem Cell Transplantation. Biol Blood Marrow Transplant. 2020.

7. Harris B, Morjaria SM, Littmann ER, et al. Gut Microbiota Predict Pulmonary Infiltrates after Allogeneic Hematopoietic Cell Transplantation. Am J Respir Crit Care Med. 2016;194(4):450–463.

8. Bartelink IH, Belitser SV, Knibbe CA, et al. Immune reconstitution kinetics as an early predictor for mortality using various hematopoietic stem cell sources in children. Biol Blood Marrow Transplant. 2013;19(2):305–313.

9. Peled JU, Devlin SM, Staffas A, et al. Intestinal Microbiota and Relapse After Hematopoietic-Cell Transplantation. J Clin Oncol. 2017;35(15):1650–1659.

10. Taur Y, Jenq RR, Perales MA, et al. The effects of intestinal tract bacterial diversity on mortality following allogeneic hematopoietic stem cell transplantation. Blood. 2014;124(7):1174–1182.

11. Holler E, Butzhammer P, Schmid K, et al. Metagenomic analysis of the stool microbiome in patients receiving allogeneic stem cell transplantation: loss of diversity is associated with use of systemic antibiotics and more pronounced in gastrointestinal graft-versus-host disease. Biol Blood Marrow Transplant. 2014;20(5):640–645.

12. Peled JU, Gomes ALC, Devlin SM, et al. Microbiota as Predictor of Mortality in Allogeneic Hematopoietic-Cell Transplantation. N Engl J Med. 2020;382(9):822–834.

13. Masetti R, Zama D, Leardini D, et al. The gut microbiome in pediatric patients undergoing allogeneic hematopoietic stem cell transplantation. Pediatr Blood Cancer. 2020:e28711.

14. Willing BP, Russell SL, Finlay BB. Shifting the balance: antibiotic effects on host-microbiota mutualism. Nat Rev Microbiol. 2011;9(4):233–243.

15. Venkataraman A, Sieber JR, Schmidt AW, Waldron C, Theis KR, Schmidt TM. Variable responses of human microbiomes to dietary supplementation with resistant starch. Microbiome. 2016;4(1):33.

16. Biagi E, Zama D, Rampelli S, et al. Early gut microbiota signature of aGvHD in children given allogeneic hematopoietic cell transplantation for hematological disorders. BMC Med Genomics. 2019;12(1):49.

17. Ingham AC, Kielsen K, Cilieborg MS, et al. Specific gut microbiome members are associated with distinct immune markers in pediatric allogeneic hematopoietic stem cell transplantation. Microbiome. 2019;7(1):131.

18. Shono Y, Docampo MD, Peled JU, et al. Increased GVHD-related mortality with broad-spectrum antibiotic use after allogeneic hematopoietic stem cell transplantation in human patients and mice. Sci Transl Med. 2016;8(339):339ra371.

19. Bilinski J, Robak K, Peric Z, et al. Impact of Gut Colonization by Antibiotic-Resistant Bacteria on the Outcomes of Allogeneic Hematopoietic Stem Cell Transplantation: A Retrospective, Single-Center Study. Biol Blood Marrow Transplant. 2016;22(6):1087–1093.

20. Yoshioka K, Kakihana K, Doki N, Ohashi K. Gut microbiota and acute graft-versus-host disease. Pharmacol Res. 2017;122:90–95.

21. Liu C, Frank DN, Horch M, et al. Associations between acute gastrointestinal GvHD and the baseline gut microbiota of allogeneic hematopoietic stem cell transplant recipients and donors. Bone Marrow Transplant. 2017;52(12):1643–1650.

22. Reikvam H, Hatfield K, Bruserud O. The pretransplant systemic metabolic profile reflects a risk of acute graft versus host disease after allogeneic stem cell transplantation. Metabolomics. 2016;12(1):12.

23. Haak BW, Littmann ER, Chaubard JL, et al. Impact of gut colonization with butyrate-producing microbiota on respiratory viral infection following allo-HCT. Blood. 2018;131(26):2978–2986.

24. Markey KA, Schluter J, Gomes ALC, et al. The microbe-derived short-chain fatty acids butyrate and propionate are associated with protection from chronic GVHD. Blood. 2020;136(1):130–136.

25. Romick-Rosendale LE, Haslam DB, Lane A, et al. Antibiotic Exposure and Reduced Short Chain Fatty Acid Production after Hematopoietic Stem Cell Transplant. Biol Blood Marrow Transplant. 2018;24(12):2418–2424.

26. Lewis JD, Chen EZ, Baldassano RN, et al. Inflammation, Antibiotics, and Diet as Environmental Stressors of the Gut Microbiome in Pediatric Crohn ‘s Disease. Cell Host Microbe. 2015;18(4):489–500.

27. Wu GD, Chen J, Hoffmann C, et al. Linking long-term dietary patterns with gut microbial enterotypes. Science. 2011;334(6052):105–108.

28. Clarke EL, Taylor LJ, Zhao C, et al. Sunbeam: an extensible pipeline for analyzing metagenomic sequencing experiments. Microbiome. 2019;7(1):46.

29. Wood DE, Salzberg SL. Kraken: ultrafast metagenomic sequence classification using exact alignments. Genome Biol. 2014;15(3):R46.

30. Kanehisa M, Furumichi M, Tanabe M, Sato Y, Morishima K. KEGG: new perspectives on genomes, pathways, diseases and drugs. Nucleic Acids Res. 2017;45(D1):D353–D361.

31. Segata N, Waldron L, Ballarini A, Narasimhan V, Jousson O, Huttenhower C. Metagenomic microbial community profiling using unique clade-specific marker genes. Nat Methods. 2012;9(8):811–814.

32. Vital M, Howe AC, Tiedje JM. Revealing the bacterial butyrate synthesis pathways by analyzing (meta)genomic data. mBio. 2014;5(2):e00889.

33. Evans CR, Karnovsky A, Kovach MA, Standiford TJ, Burant CF, Stringer KA. Untargeted LC-MS metabolomics of bronchoalveolar lavage fluid differentiates acute respiratory distress syndrome from health. J Proteome Res. 2014;13(2):640–649.

34. Weber D, Jenq RR, Peled JU, et al. Microbiota Disruption Induced by Early Use of Broad-Spectrum Antibiotics Is an Independent Risk Factor of Outcome after Allogeneic Stem Cell Transplantation. Biol Blood Marrow Transplant. 2017;23(5):845–852.

35. Galloway-Pena JR, Peterson CB, Malik F, et al. Fecal Microbiome, Metabolites, and Stem Cell Transplant Outcomes: A Single-Center Pilot Study. Open Forum Infect Dis. 2019;6(5):ofz173.

36. Lahteenmaki K, Wacklin P, Taskinen M, et al. Haematopoietic stem cell transplantation induces severe dysbiosis in intestinal microbiota of paediatric ALL patients. Bone Marrow Transplant. 2017;52(10):1479–1482.

37. Zaneveld JR, McMinds R, Vega Thurber R. Stress and stability: applying the Anna Karenina principle to animal microbiomes. Nat Microbiol. 2017;2:17121.

38. Dethlefsen L, Relman DA. Incomplete recovery and individualized responses of the human distal gut microbiota to repeated antibiotic perturbation. Proc Natl Acad Sci U S A. 2011;108 Suppl 1:4554–4561.

39. Becattini S, Taur Y, Pamer EG. Antibiotic-Induced Changes in the Intestinal Microbiota and Disease. Trends Mol Med. 2016;22(6):458–478.

40. Jakobsson HE, Jernberg C, Andersson AF, Sjolund-Karlsson M, Jansson JK, Engstrand L. Short-term antibiotic treatment has differing long-term impacts on the human throat and gut microbiome. PLoS One. 2010;5(3):e9836.

41. Galloway-Pena JR, Kontoyiannis DP. The gut mycobiome: The overlooked constituent of clinical outcomes and treatment complications in patients with cancer and other immunosuppressive conditions. PLoS Pathog. 2020;16(4):e1008353.

42. Vemuri R, Shankar EM, Chieppa M, Eri R, Kavanagh K. Beyond Just Bacteria: Functional Biomes in the Gut Ecosystem Including Virome, Mycobiome, Archaeome and Helminths. Microorganisms. 2020;8(4).

43. Singh RK, Chang HW, Yan D, et al. Influence of diet on the gut microbiome and implications for human health. J Transl Med. 2017;15(1):73.

44. Ziegler M, Han JH, Landsburg D, et al. Impact of Levofloxacin for the Prophylaxis of Bloodstream Infection on the Gut Microbiome in Patients With Hematologic Malignancy. Open Forum Infect Dis. 2019;6(7):ofz252.

45. Mathewson ND, Jenq R, Mathew AV, et al. Gut microbiome-derived metabolites modulate intestinal epithelial cell damage and mitigate graft-versus-host disease. Nat Immunol. 2016;17(5):505–513.

46. Shono Y, Docampo MD, Peled JU, et al. Increased GVHD-related mortality with broad-spectrum antibiotic use after allogeneic hematopoietic stem cell transplantation in human patients and mice. Sci Transl Med. 2016;8(339):339ra371.

47. Zhu C, Fuchs CD, Halilbasic E, Trauner M. Bile acids in regulation of inflammation and immunity: friend or foe? Clin Exp Rheumatol. 2016;34(4 Suppl 98):25–31.

48. Masetti R, Zama D, Leardini D, et al. Microbiome-Derived Metabolites in Allogeneic Hematopoietic Stem Cell Transplantation. Int J Mol Sci. 2021;22(3).

49. Joshi NM, Hassan S, Jasani P, et al. Bile acid malabsorption in patients with graft-versus-host disease of the gastrointestinal tract. Br J Haematol. 2012;157(3):403–407.

50. Simms-Waldrip TR, Sunkersett G, Coughlin LA, et al. Antibiotic-Induced Depletion of Anti-inflammatory Clostridia Is Associated with the Development of Graft-versus-Host Disease in Pediatric Stem Cell Transplantation Patients. Biol Blood Marrow Transplant. 2017;23(5):820–829.

