## Supplemental material for "Early microbiome and metabolome signatures in pediatric patients undergoing allogeneic hematopoietic cell transplantation"

SUPPLEMENTAL TABLE

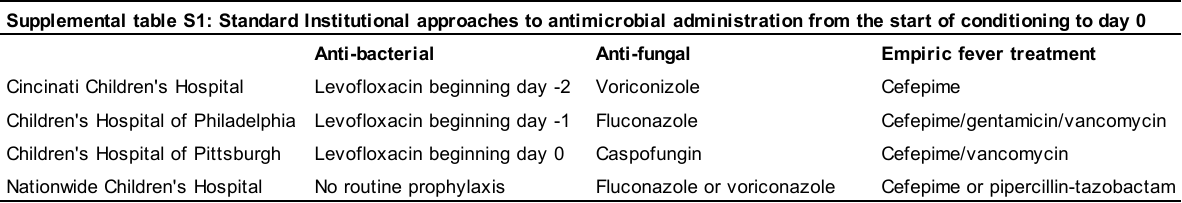

SUPPLEMENTAL FIGURE S1: Heatmap with relative abundance of bacteria taxa at genus level.

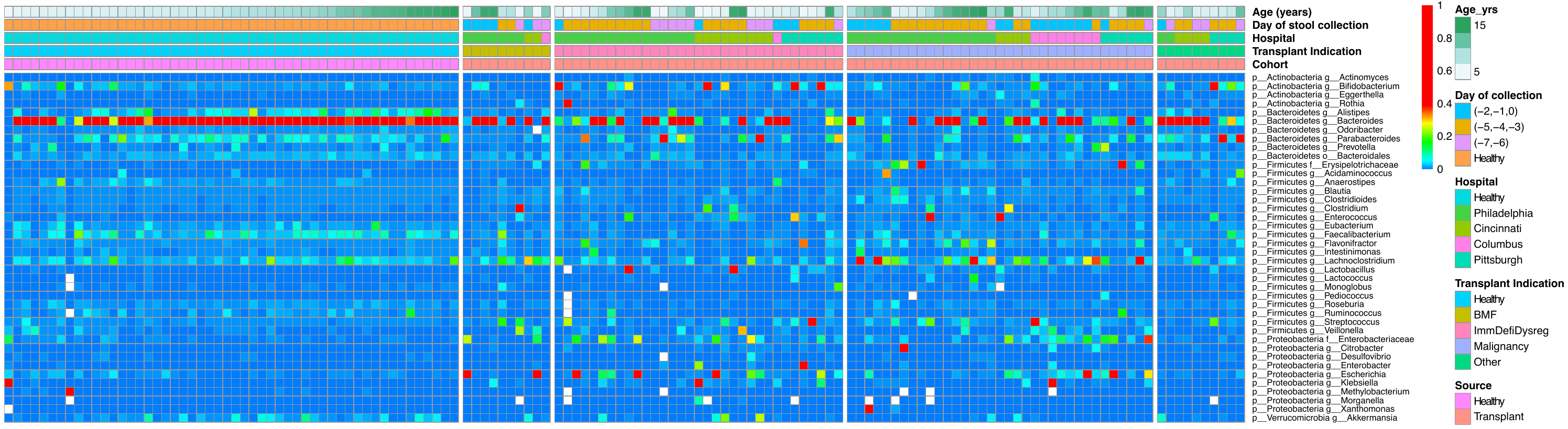

SUPPLEMENTAL FIGURE S2: Heatmap with gene abundance. 7,658 KEGG orthology items (K numbers) are found in the data. 73 K numbers whose relative abundance is at least 0.5% in at least one sample are included below and shown as the square root transformation of relative abundance.

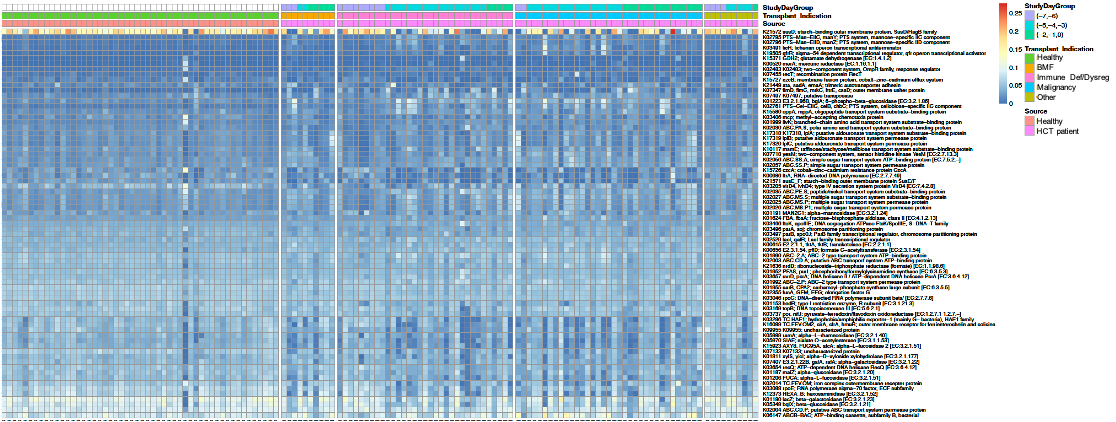
